# Individual participant data systematic reviews with meta-analyses of psychotherapies for borderline personality disorder: A protocol

**DOI:** 10.1101/2020.11.27.20238394

**Authors:** Ole Jakob Storebø, Johanne Pereira Ribeiro, Mickey T. Kongerslev, Jutta Stoffers-Winterling, Mie Sedoc Jørgensen, Klaus Lieb, Anthony Bateman, Richard Kirubakaran, Nicolas Dérian, Eirini Karyotaki, Pim Cuijpers, Erik Simonsen

## Abstract

**Introduction:** The heterogeneity in people with BPD and the range of specialised psychotherapies means that people with certain BPD characteristics might benefit more or less from different types of psychotherapy. Identifying moderating characteristics of individuals is a key to refine and tailor standard treatments so they match the specificities of the individual patient. The objective of this is to improve the quality of care and the individual outcomes. Thus, the aim of the current reviews is to investigate potential predictors and moderating patient characteristics on treatment outcomes for patients with BPD.

**Methods and analysis:** Our primary meta-analytic method will be the one-stage random-effects approach. To identify predictors, we will be using the one-stage model that accounts for interaction between covariates and treatment allocation. Heterogeneity in case-mix will be assessed using a membership model based on a multinomial logistic regression where study membership is the outcome. A random-effects meta-analysis is chosen to account for expected levels of heterogeneity.

**Ethics and dissemination:** The statistical analyses will be conducted on anonymised data that have already been approved by the respective ethical committees that originally assessed the included trials. The three IPD reviews will be published in high impact factor journals and their results will be presented at international conferences and national seminars.

**Protocol registration:** The IPD reviews, described in this study protocol, are registered with the PROSPERO International Prospective Register of Systematic Reviews (registration number: awaiting)

**Strengths and limitations of this protocol:**

- These IPD-reviews are the first to systematically review and investigate psychotherapy for people with borderline personality disorder using individual participant data.
- The IPD-reviews will provide information on moderators and predictors in patients with borderline personality disorder that predict who may benefit most from which type of specialised psychotherapy.
- Individual participant data allows for a more precise risk of bias assessment and decreases the amount of unclear risk of bias in many of the included trials.
- A limitation to IPD-reviews in general is that data retrieval can be challenging.
- The IPD-reviews are limited to the outcomes and patient characteristics that have been assessed in the included trials.

## Introduction

### Borderline personality disorder - diagnosis and treatment

Due to the polythetic approach to diagnosing borderline personality disorder (BPD), there are 256 ways of meeting the criteria for a BPD diagnosis.^1^ This means that apart from meeting the general diagnostic criteria for personality disorder (PD), the patients also need to fulfil five or more of the nine specific BPD criteria according to the current DSM classification system.^1^ This makes the BPD population highly heterogeneous. A fact that is exacerbated by the common co-occurrence of many other psychiatric and somatic conditions. Also, co-occurring psychiatric conditions, e.g., life-threatening eating disorders or substance use dependence, are often persistent and may impede BPD treatment.^2-4^ People with BPD need effective treatment due to the considerable psychological suffering of those concerned^5^, the high burden experienced by their families and significant others,^6 7^ the significant impact they have on mental health services given their frequent use,^8 9^ as well as the association of BPD with sustained functional impairment,^10^ physical illness,^11^ and premature death.^12-14^

The prevalence of BPD in the general population is estimated to be 1,8%,^15^ and the diagnosis is one of the most common in the psychiatric system.^16^ In addition to the effects on the individual and his/hers relatives, the annual direct total costs for the Danish health sector is roughly 53.000 Euro per patient with BPD per year. This number is 16 times the costs of general population controls, according to a recent nationwide study by Hastrup and colleagues.^17^ From an economic perspective alone, this calls for more effective treatments for people with BPD, and a precisely targeted use of resources.

Storebø and colleagues published a Cochrane Review in May 2020^18^ that investigated the beneficial and harmful effects of psychotherapeutic treatments for people with BPD. Their findings showed that all major types of psychotherapies for BPD had a modest positive average effect at group level. However, it is likely that the participants’ individual responses differed in relation to certain self-inherent characteristics. Therefore, data is now needed at the level of the individual patient in order to find out for *whom* the different specialised psychotherapies may have a greater or smaller effect (i.e., what type of psychotherapy will have the largest treatment effect when taking the personal and clinical characteristics of the participant into consideration).

Given the heterogeneity of individuals affected by BPD, and the availability of several effective treatments of different theoretical orientations^18^ it is possibly that individuals with certain characteristics might benefit to a higher extent from some treatments, and less from others.

Identifying such patient characteristics may allow for a more refined and individualised treatment, and optimise the treatment quality and effect for BPD patients.^19^ Research identifying BPD characteristics that affect the outcome of the various treatments is therefore needed.

As called for by Barber and Solomov,^20^ we attempt to find and match the most effective specialised psychotherapeutic treatments with the needs of the individual patient based on personal and clinical characteristics. Hereby, we are effectively moving towards a personalised approach to psychotherapeutic treatment. Most people with BPD will receive psychological interventions because drugs are not effective for the BPD core symptoms.^4 21 22^ Psychological interventions are often provided for relatively long periods (e.g., one year or longer).^23 24^ Psychotherapy is thus the current treatment of choice for patients with BPD.^25^

A broad range of specialised psychotherapies for BPD are available.^18 26 27^ These therapies are usually precisely structured and manualised^26^ and are delivered in individual therapy format or as combined individual- and group treatments. Most BPD-specific psychological interventions involve multimodal therapy, treatment contracts, actively taking measures to minimise premature non-completion of treatment, providing a crisis intervention protocol and stimulating the participant’s sense of agency.^26-31^ For a more detailed description of the different types of BPD tailored psychotherapies, see Storebø et al. 2020.^18^

Most people in treatment for BPD receive long-term psychotherapeutic treatment,^4 24^ while, on the other hand, not all individuals in need have access to adequate treatment, even in highly-developed countries.^32^ A recent review of European guidelines on diagnosing and treating personality disorders reported that psychotherapy was the first-line treatment recommended in all countries.^25^

Psychotherapeutic treatments for BPD are based on a variety of different therapeutic schools, e.g., psychodynamic, cognitive-behavioural, or client-centered/humanistic therapy.^33^ However, there has been a development of multiple psychotherapeutic treatments that are more disorder-specific (i.e. specifically adapted for BPD) within the last three decades. This development is due to the disorder-inherent challenges that individuals diagnosed with BPD often face and pose in treatment. Among the specific psychological interventions for people diagnosed with BPD, the most commonly researched and used ones are: Dialectical behaviour therapy (DBT),^34 35^ Mentalisation-based treatment (MBT),^36-38^ Systems training for emotional predictability and problem solving (STEPPS),^39^ Transference-focused therapy (TFP), ^40 41^ Cognitive analytic therapy (CAT),^42 43^ and Schema-focused therapy (SFT).^44 45^ The treatment of BPD is very complex due to the complexity of the pathology itself, but tailored treatments can improve the outcomes. Therefore we want to systematize the use of treatments matching patients characteristics by conducting these IPD reviews.

### Purpose of the current Individual Patient Data reviews

The preceding Cochrane review of psychological psychotherapies for BPD^18^ provided an initial overview of the research in the area and presented results based on analyses of aggregated data. As such, this Cochrane review can be considered a first step in the research process. The current project is the next steps which focus on predictors and moderators of outcomes. ^46^

We define predictors as a collection of parameters (demographic, clinical or biologic) that influence the likelihood of specific outcomes to occur.

Moderators are special case of predictors defined as baseline parameters (demographic, clinical or biologic) affecting the likelihood of a specific event to occur in one situation compare to another one. E.g., a mediator can improve the prediction of a treatment efficacy to achieve a specific outcome, compare to another treatment^47^

The results of the project will provide tangible advice to practitioners and people affected by the disorder on how to select the psychotherapeutic treatment deemed to have the most effective outcome when considering patient characteristics. Overall, this will help to ensure that more people with BPD will receive a treatment that is adapted to the individual’s needs. To investigate these characteristics, we will perform three systematic reviews with meta-analyses of psychotherapies for BPD using individual participant data (IPD). IPD meta-analyses are particularly well suited for the purposes of this project because all the raw data from the included trials is used, which allows for a detailed exploration of the causes of heterogeneity.^48^ IPD reviews are closely related to personalised medicine where it is important to understand for whom, and under what conditions, treatment exerts the best effect. Furthermore, findings of the reviews are likely to inform future treatment guidelines.

### IPD review methodology

Though the IPD methodology is still rather new, IPD reviews have generally had a substantial impact on clinical practice and research.^46^

When IPD for each participant in clinical trials are available, they can be used to individualise the results of clinical trials.^26^ There are already several examples of recent IPD reviews that has decreased the knowledge-gap in somatic research areas.^49-51^ Within the psychiatric field, IPD reviews have been used to investigate treatment effects across various patient groups, with direct implications for clinical practice.^52-57^ However, when conducting extensive searches in relevant databases, we found no IPD review that investigated psychotherapy for BPD.

The use of IPD can promote standardisation of data in analyses and allows for direct extraction of data to outcomes, independently of how these were reported in the original trial publication. Studies that use IPD show a greater power in detecting effect differences in outcomes between individuals.^58^ This can provide valuable information about responders and non-responders to the different types of treatments. Analyses based on IPD data also allow for the use of more sophisticated statistical methods.^59^ In particular, IPD may allow for exploring causes of heterogeneity such as baseline differences, selection criteria, dose and duration of treatments received by participants in control groups, and differential negative effects of the treatments. Missing data can also be handled in a more standardised manner in IPD reviews. Furthermore, access to IPD data allows for a more reliable risk of bias assessment due to deeper insight into the original data. Finally, IPD allows us to perform subgroup analyses that have not previously been conducted, thereby answering new and pressing research questions concerning how to optimise treatments for BPD for the individual patient.^48^

### Objectives

This protocol describes three planned IPD reviews each aiming to answer different salient research questions that remain pertinent based on prior literature, and especially the recently published Cochrane review on the topic ^18^:

#### IPD review 1: BPD symptom severity and interpersonal functioning

1.1) What are the effects of different psychotherapies when compared with unspecific controls (e.g., treatment usual (TAU), wait-list (WL) or no-intervention (NI) and specific psychotherapeutic interventions for people with BPD on the primary outcomes: BPD symptom severity and interpersonal functioning?

1.2) What are the moderators of the differential efficacy between psychotherapy versus control conditions in reducing BPD symptom severity and increasing interpersonal functioning?

1.3) What are the prognostic factors and effect moderators associated with the secondary outcomes: Serious and non-serious adverse events.

#### IPD review 2: Quality of life and psychosocial functioning

2.1) What are the effects of different psychotherapies when compared with unspecific controls (e.g., TAU, WL or NI) and specific psychotherapeutic interventions for people with BPD on the primary outcomes quality of life and psychosocial functioning?

2.2) What are the moderators of the differential efficacy between psychotherapy versus controls in quality of life and psychosocial functioning?

2.3) What are the prognostic factors and effect moderators associated with the secondary outcomes: Serious and non-serious adverse events?

#### IPD review 3: Suicidality and self-harm

3.1) What are the effects of different psychotherapies when compared with unspecific controls (e.g., TAU, WL or NI) and specific psychotherapeutic interventions for people with BPD on the primary outcomes: Suicidality and self-harm?

3.2) What are the moderators of the differential efficacy between psychotherapy versus controls in reducing suicidality and non-suicidal risk behaviour?

3.3) What are the prognostic factors and effect moderators associated with the secondary outcomes: Serious and non-serious adverse events?

## Method and analysis

### General approach

The current protocol follows the general guidance provided as part of the Preferred Reporting Items for Systematic Reviews and Meta-Analyses (PRISMA)-IPD statement^60^ (see checklist S1 in supplemental material).

### Search criteria

To meet our inclusion criteria, at least 70% of participants in a trial are required to have a formal diagnosis of BPD according to DSM-III-R and onwards.^1^ We will include trials with subsamples of people with BPD when data is provided separately on BPD participants. We will not include trials that focus on people with mental impairment, organic brain disorder, dementia or other severe neurologic/neurodevelopmental diseases or people with medical health issues, e.g., cancer or HIV.

The search will not be limited by language, year of publication or type of publication. We will seek translation of relevant sections of articles that are not in English.

### Search method for identification of studies

Our search strategy for eligible studies will be based on the searches conducted in the prior Cochrane review on psychological therapies for BPD.^18^ These searches will be updated with a top-up search which is described in detail below (see supplemental material S2 for search string).

### Databases

We will search for eligible studies in the following 22 databases and registries: Cochrane Central Register of Controlled Trials, MEDLINE Ovid, Embase Ovid, CINAHL EBSCOhost, PsycINFO Ovid, ERIC EBSCOhost, BIOSIS Previews, Web of Science Core Collection Clarivate Analytics, Sociological Abstracts ProQuest, LILACS, OpenGrey, JISC Library Hub Discover (previously COPAC), Proquest Dissertations and Theses Global, DART Europe E-Theses Portal, Networked Digital Library of Theses and Dissertations (NDLTD), Australian New Zealand Clinical Trials Registry, Clinicaltrials.gov, EU Clinical Trials Register, Open Trials, ISRCTN Registry, Be Part of Research, WHO International Clinical Trials Registry Platform (ICTRP).

### Types of studies

The studies that will be included in our search are randomised clinical trials (RCTs) that compare psychotherapeutic treatments for BPD with unspecific controls (e.g., TAU, WL, and NI) and specific psychotherapeutic treatments.

### Population

The studies will include people of all ages, any gender, in any setting, with a formal, categorical diagnosis of BPD according to the Diagnostic and Statistical Manual of Mental Disorders (DSM) Third Edition (DSM-III; APA 1980), Third Edition Revised (DSM-III-R; APA 1987), Fourth Edition (DSM-IV; APA 1994), Fourth Edition Text Revision (DSM-IV-TR; APA 2000), and Fifth Edition (DSM-5; APA 2013), or the Emotionally unstable personality disorder, borderline type in International Classification of Diseases and Related Health Problems (ICD) 10th version (WHO 1993), with or without comorbid conditions.^18^

### Intervention

We will search for well-defined theory driven psychological interventions regardless of theoretical orientation (e.g., psychodynamic therapy, cognitive behavioural therapy, systemic therapy or eclectic therapies designed for BPD treatment), in any kind of treatment setting (e.g., inpatient, outpatient or day clinic) and mode (individual, group or combined therapy),

### Study selection

The paper titles and abstract identified in the top-up search will be independently screened by two members of the project group to remove those that are clearly ineligible. Similarly, two reviewers will read the full text articles independently. Disagreements about study inclusion will be resolved by discussion with a third review author. All trials excluded from the review after the full text level will be given reasons for exclusion.

### Quality assessment

Study quality will be assessed by two reviewers from the project group who will independently evaluate the studies using the updated Cochrane Risk of Bias tool (RoB 2) in the quality assessment of included studies.^61^

Studies will be rated on each criterion with either ‘low risk’, ‘high risk’ or ‘some concerns’. Each study as a whole will be rated according to its highest risk of bias in any of the assessed domains. i.e., if any domain is judged as having a high risk of bias, the whole study will be classified as “high risk of bias”. We will assess the following domains: 1) bias arising from the randomisation process, 2) bias due do deviations from the intended interventions, 3) bias due to missing outcome data, 4) bias due to measurement of the outcome, and 5) bias due to selective reporting.^59^

### Data collection process

To be able to get raw data from the included RCTs, we will obtain contact information through the included publications or by an online search. We will contact the authors of each included RCT and provide them with the IPD review protocol and a cover letter explaining what the study is about. If we receive no response, we will send a reminder after one week and again after one month before excluding the trial for unavailability.

### IPD-BPD consortium

All RCT authors will be invited to be part of an IPD-BPD consortium that the project group will establish. The name of this consortium will be “IPD-BPD”. The aim of this taskforce is to support the project, make it easier to have authors participate, to increase awareness within the public and clinical community, and to help with dissemination of results. All RCT authors will be invited to be co-authors of the IPD reviews.

### Developing the IPD-BPD database

IPD will be extracted from all included RCTs where the authors are willing to share their data. The IPD will be exported and integrated into a spreadsheet. A template spreadsheet will be created and pilot-tested. We will need data from all randomised patients (intention-to-treat samples) of all included trials. We will make a list of variables that we need and send this to the authors of the included trials. Furthermore, we will ask for the formal data codes and time points at which data was collected.

Raw data (de-identified data) can be transferred by a secure electronic transfer. The data sent from authors will be checked for completeness and accuracy. We will compare the participant numbers, descriptive data and outcome data to the reported data in the original peer-reviewed article. If any irregularity is present, the issue will be discussed with the study authors for clarification. Raw datasets will be saved in their original formats and then exported into a common format. The data will be stored on a secure server. We will rename the variables for each study in a consistent manner. All individual datasets will be merged into one large IPD dataset that takes the study clusters into account.^62^

### Data items

#### Primary outcomes

Primary outcomes will be measured by the use of standardised psychometric rating scales. We will included both self-rated and observer-rated measures. If both are available we will prefer observer-rated.

#### IPD review 1

1.1 BPD symptom severity, e.g., assessed by the Zanarini Rating Scale for Borderline Personality Disorder (Zan BPD) ^63^, the Borderline Personality Disorder Severity Index, Fourth version (BPDSI-IV) ^64^, or the Clinical Global Impression Scale for people with Borderline Personality Disorder (CGI-BPD).^65^

1.2 Interpersonal functioning, assessed by, for example, the Inventory of Interpersonal Problems^66^ (IIP), or the relevant item or subscale on the Zan BPD, ^63^ CGI BPD,^65^ - - BPDSI IV. ^64^

#### IPD review 2

2.1 Quality of life, e.g., assessed by the The Quality of Life Satisfaction and Enjoyment^67^ or the EuroQol five-dimensional.^68^

2.2 Psychosocial functioning, e.g., assessed by the Global Assessment Scale^69^, the Global Assessment of Functioning Scale^70^, or the Social Functioning Questionnaire.^71^

#### IPD review 3

3.1 Self-harm, in terms of the proportion of participants with self-harming behaviour, or assessed by e.g., the Deliberate Self-harm Inventory^72^ or the Self harm behaviour Questionnaire.^73^

3.2 Suicide-related outcomes, e.g., assessed by the Suicidal Behaviours Questionnaire^74^ or the Beck Scale for Suicidal Ideation^75^, or in terms of the proportion of participants with suicidal acts.

### Secondary outcomes

Adverse effects will be measured by the use of standardised psychometric rating scales, such as the Systematic Assessment for Treatment Emergent Events,^76^ by laboratory values or spontaneous reporting. We will divide the reported adverse effects into severe and non-severe, according to the International Committee of Harmonization guidelines.^77^ We will define serious adverse effects as any event that led to death, was life-threatening, required inpatient hospitalisation or prolongation of existing hospitalisation, resulted in persistent or significant disability, or any important medical event that may have jeopardised the participant’s health or required intervention to prevent one of the aforementioned outcomes occurring. We will consider all other adverse effects to be non-serious. Additionally, deterioration will be examined.

### Effect predictors and moderators

We want to know which patient characteristics predict a reduction of the primary outcomes for the three IPD reviews: IPD review 1: BPD symptom severity and interpersonal functioning. IPD review 2: Quality of life and psychosocial functioning, IPD review 3: Self-harm, suicidal behaviour, and regardless of treatment allocation (predictor variables).

In three IPD reviews, we aim to investigate which patient characteristics predict treatment response in terms of three sets of outcomes: See figure 1.

**Figure 1:**
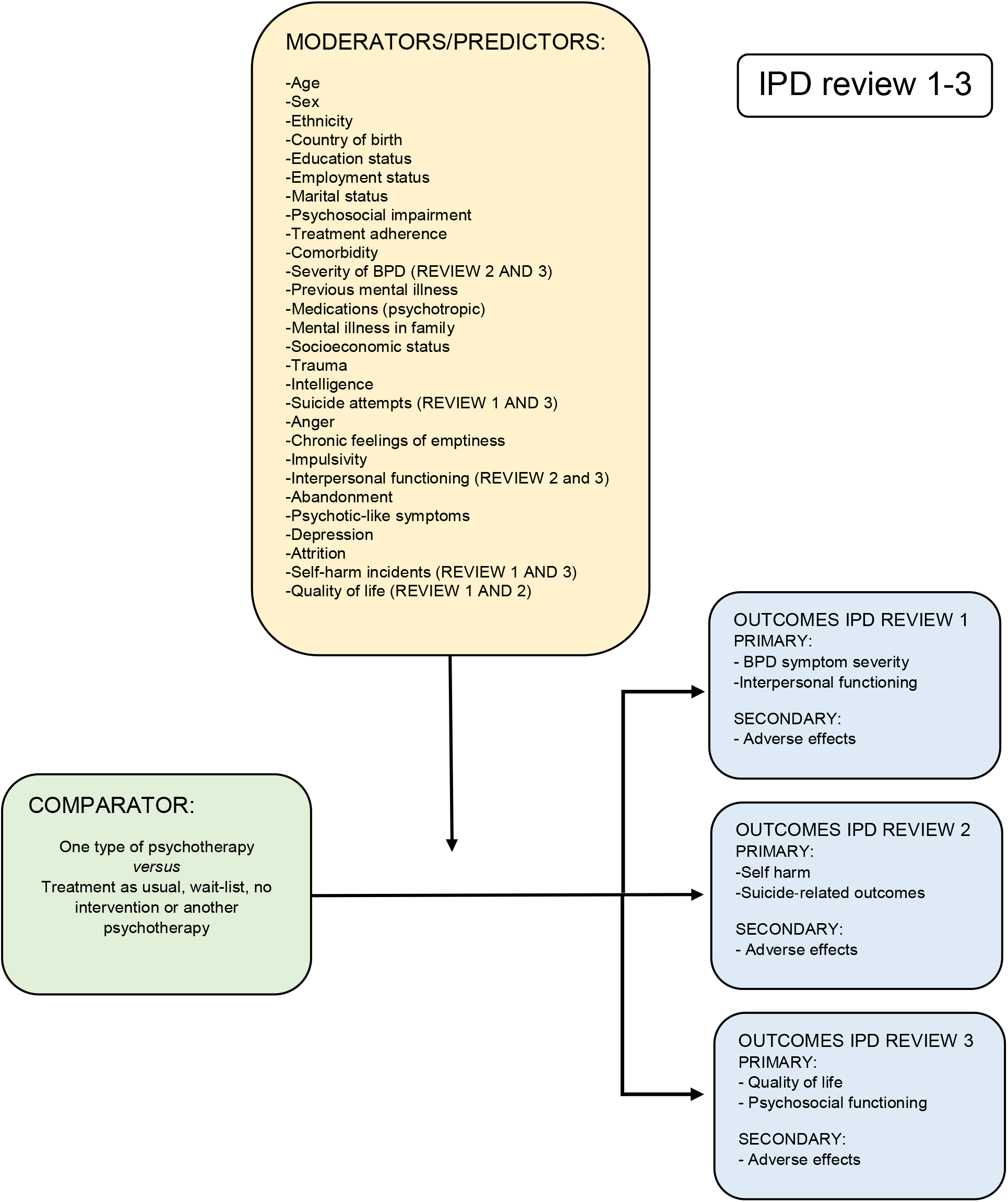
Comparisons, moderators and outcomes in IPD review 1-3.

We also intend to identify moderators, i.e., variables which affect outcomes based on treatment allocation.^78^ Moderators differentiate between the effects of two treatments and predictors refer to prognostic factors.^78^

Patient characteristics will be included in the analyses, if they are consistently reported, available across datasets and justify inclusion based on prior literature that identifies them as potential predictors or moderators.^78^ Such characteristics could be age at baseline, sex, ethnicity, country of birth, education status, employment status, marital status, severity of BPD, psychosocial impairment, treatment adherence, comorbidity, previous mental illness, medications (psychotropic), mental illness in family, socioeconomic status, trauma, IQ, suicide attempts, anger, chronic feelings of emptiness, impulsivity, interpersonal problems, abandonment, psychotic-like symptoms, depression and self-harm incidents. We will examine the published papers and verify which moderators are investigated. We will include all moderators that are investigated in at least two studies.

### Data analysis

Our primary meta-analytic method will be the one-stage random-effects approach, which is particularly suitable for investigating predictors and moderators compared with the two-stage method. The one-stage random-effects method is also less influenced by the expected small size of some the studies included in the planned meta-analyses.^79^

To identify predictors, we will use the one-stage model that accounts for interaction between covariates and treatment allocation. Covariates with statistical evidence for association with the outcome will be added in a unique global model. Significant association (p < 0.05) with the outcome in the global model will then be used to identify the predictors. Similarly, we will use a one-stage approach to identify moderators by investigating the interaction between selected covariates and the treatments, one covariate at the time.^78^ To account for potential ecological bias, covariates will be transformed at study-level before analysis using the proper methodology. ^80^

Datasets will be checked for their completeness and integrity. To handle missing values, we will use multiple imputation under the missing at random assumption.^81^ Missing data will be imputed within each original study before data of the individual studies are pooled. A sensitivity analysis will be conducted using a pattern-mixture approach.^82^

Heterogeneity in case-mix will be assessed using a membership model based on a multinomial logistic regression where study membership is the outcome. The derived c-statistics will reflect the difference in baseline characteristics and outcome.^83^ As a certain level of heterogeneity is expected (e.g., due to differences in study populations, types of psychotherapy, or differences in the control group) a random-effects meta-analysis is chosen to account for these variations.

All analyses will be conducted using a well-established statistical platform providing ready-to-use packages and libraries to perform such analyses, like STATA.^84^

### Subgroup analyses

In addition, we will perform meta-analyses including only studies classified as ‘low risk’ of bias to assess the impact of studies of lower methodological quality and type of control conditions on the findings. When possible, a similar approach will be used to compare studies based on differences in the criteria for the risk of bias.^85^

### Difference between included and not-included studies in the IPD review

We will compare the dataset on the primary outcomes from the previously published Cochrane review ^18^ with the data included in the present IPD reviews. If there is a discrepancy between the datasets, we will report both results. If necessary (depending on the outcome of subgroup analyses), we will execute the appropriate approach of combining the aggregated data and the IPD data to perform either: Meta-analyses of the aggregated data, meta-analyses of reconstructed IPD or hierarchical-related regressions.^86^

### Further development of the analysis plan

We will write a more detailed plan for the statistical analyses in the period from receiving the data to the actual data analyses. In that plan, we will specify how covariates will be modelled (i.e., whether quantitative patient-level characteristics such as age is treated as continuous or categorical).

### Patient and public involvement

We are collaborating with three Danish patient- and family alliance organisations addressing BPD and mental illness. Representatives from all three organisations have read and commented on the protocol. We are taking this approach to keep the project anchored and in proximity to clinical practice. Hereby we indirectly give means to individuals with BPD to influence the research process.

We will similarly invite the members of the patient and family alliance organisations to comment on the IPD reviews before publishing them. We do so to offer a sense of ownership and inclusion in the project.

## Supporting information

S1 - Search string

S2 - PRISMA IPD checklist

## Data Availability

This is a protocol of three systematic reviews which does not contain any data to available.

## Ethics and dissemination

### Ethics approval and consent to participate

Approval by a research ethics committee is not required to conduct these reviews because they involves statistical analyses of anonymous data that have already been approved by the respective ethical committees originally assessing the included trials.

### Publications

The three IPD reviews will be published in high impact factor journals. The results from the reviews will be presented at international conferences as well as in national seminars and conferences.

## Authors’ contributions

All authors contributed to writing this protocol.

## Competing interests

Ole Jakob Storebø: Trained in child and adolescent psychoanalytic play therapy and trained in group psychoanalysis.

Johanne Pereira Ribeiro: No competing interests.

Mickey T. Kongerslev: Trained in mentalisation-based and psychodynamic psychotherapy, and conducts research and training in mentalisation-based therapy. Has written books on mentalisation-based therapy.

Jutta Stoffers-Winterling: Board-certified behaviour therapist, trained in dialectical behaviour therapy.

Mie Sedoc Jørgensen: Associated with the M-GAB trial, trained in dialectical behaviour therapy and psychodynamic therapy.

Klaus Lieb: Board-certified cognitive behaviour therapist with a special interest in schema therapy. KL has been involved in trials investigating inpatient dialectical behaviour therapy (Bohus 2004); and inpatient schema focused therapy (Reiss 2014).

Anthony Bateman: Receives honoraria for training in mentalisation-based treatment for BPD.

Nicolas Dérian: No competing interests. Richard Kirubakaran: No competing interests. Eirini Karyotaki: No competing interests.

Pim Cuijpers: No competing interests.

Erik Simonsen: PI of the M-GAB trial, trained in group psychoanalysis.

