## Supplementary material for "Individual participant data systematic reviews with meta-analyses of psychotherapies for borderline personality disorder: A protocol": S1 - Search string

### S1 - Search strings from Storebø et al. 2020

**Cochrane Central Register of Controlled Trials, in the Cochrane Library**
#1 MeSH descriptor: [Borderline Personality Disorder] explode all trees
#2 borderline next state*
#3 borderline next personalit*
#4 "axis II" or "cluster B"
#5 idealization next devaluation
#6 (vulnerable or hyperbolic) next temper*
#7 (((unstab* or instab* or poor or disturb* or fail* or weak* or dysregulat*) next (self* or impuls* or interperson* or identit* or relation* or emotion* or affect*)) and (person* or character or PD))
#8 impulsiv* near personalit*
#9 (self next (injur* or damag* or destruct* or harm* or hurt* or mutilat*))
#10 suicidal next behavio?r
#11 (feel* next (empt* or bored*))
#12 (anger next control*)
#13 (risk-taking next (behavior or behaviour))
#14 #1 or #2 or #3 or #4 or #5 or #6 or #7 or #8 or #9 or #10 or #11 or #12 or #13

**Medline Ovid**
1 Borderline Personality Disorder/
2 ((borderline or border-line) adj3 (state* or personalit*)).kf,tw.
3 ("Axis II" or "Cluster B" or flamboyant or "F60.3" or "F60.30" or "F60.31").kf,tw.
4 (idealization adj5 devaluation).kf,tw.
5 ((vulnerable or hyperbolic) adj3 temperament).kf,tw.
6 (((unstab* or instab* or poor or disturb* or fail* or weak or dysregulat*) adj3 (self* or impuls* or interperson* or identit* or relationship* or emotion* or affect*)) and (personality or character or PD)).kf,tw.
7 (impulsiv* adj5 (behavio?r or character or personalit*)).kf,tw.
8 (self adj3 (injur* or damag* or destruct* or harm* or hurt* or mutilat*)).kf,tw.
9 (suicidal adj3 behavio?r).kf,tw.
10 (feel* adj3 (empt* or bored*)).kf,tw.
11 (anger adj5 control*).kf,tw.
12 (risk-taking adj3 behavio?r).kf,tw.
13 or/1-12
14 randomised controlled trial.pt.
15 controlled clinical trial.pt.
16 randomi#ed.ab.
17 placebo.ab.
18 randomly.ab.
19 trial.ab.
20 groups.ab.
21 drug therapy.fs.
22 or/14-21
23 exp Animals/ not Humans/
24 22 not 23
25 13 and 24
